# A qualitative study to explore women’s perceptions of pregnancy and antenatal care in Chizenga village, Chikwawa, Malawi

**DOI:** 10.1101/2025.10.06.25337230

**Authors:** Helen Layton, Deborah Nyirenda, Terry Kana

## Abstract

Malawi’s Demographic and Health Survey (2015–2016) reports that only 24% of women attended their first antenatal care (ANC) visit in the first trimester [1]. This study explores women’s perceptions of ANC in Chizenga village, Chikwawa, a rural setting with high maternal mortality and low ANC uptake. Using a qualitative interpretivist approach, 14 pregnant women aged 19 to 40 years were interviewed to understand their knowledge of ANC and the barriers they face. Findings show that while women value ANC and recognise its role in reducing maternal and neonatal mortality, most initiated care after the first trimester. Socioeconomic constraints—particularly distance, transport costs, and limited health literacy— contributed to delayed ANC attendance. Nevertheless, women demonstrated resilience and a strong commitment to maternal health, walking over 22 kilometres to access services. Participants voiced a need for increased ANC education and proposed community-based solutions such as health worker outreach and improved access to services. The study calls for context-sensitive policy interventions to ensure equitable access to respectful ANC for all women, regardless of socio-economic or marital status.

## Introduction

Maternal mortality remains a major global health challenge. Its significance is underscored by Sustainable Development Goal 3.1, which aims to reduce the global maternal mortality ratio (MMR) to fewer than 70 deaths per 100,000 live births by 2030 [2]. Despite ongoing efforts, over 700 women still die each day due to complications related to pregnancy and childbirth, with over 90% of these deaths occurring in low- and middle-income countries—particularly in sub-Saharan Africa. At the time of writing, this figure was estimated at 810 deaths per day, with 94% occurring in sub-Saharan Africa. [3,4,7].

Between 2000 and 2015, the global MMR declined from 342 to 223 deaths per 100,000 live births. However, since 2016, progress has plateaued in many regions [5]. As Moyer et al (2023) argue, aggregating data over long periods or across regions can obscure disparities within and between countries, especially among marginalised populations [9]. The stalling of maternal health improvements has been partly linked to the Millennium Development Goals (MDGs) era, during which efforts focused on addressing leading causes of maternal death— such as postpartum haemorrhage and sepsis—using interventions that often did not require highly specialised care. For instance, the use of oxytocin for active management of the third stage of labour became widely adopted in low-resource settings [7].

In more recent years, global crises such as the COVID-19 pandemic, climate-related disasters, economic shocks, political instability, and armed conflict have further diverted attention and resources from maternal health priorities [8]. In 2020, sub-Saharan Africa accounted for nearly 70% of all maternal deaths worldwide. In humanitarian settings, maternal mortality rates were more than double the global average—despite most deaths being preventable through timely access to quality care across the maternal health continuum [9,10].

Low utilisation of antenatal care (ANC) services is a significant contributing factor to poor maternal outcomes in low-resource countries. Women in low-income and middle-income countries are approximately 23 times more likely to die from pregnancy-related complications compared with those in high-income countries [11]. Early initiation of quality ANC—ideally within the first 12 weeks of pregnancy—is recognised as essential for reducing maternal and neonatal mortality, and for identifying conditions such as anaemia, hypertension, and infections [12].

In response to stalled progress, the World Health Organization (WHO) revised its ANC guidelines in 2016. The previous four-visit model, known as Focused Antenatal Care (FANC), was replaced with a more comprehensive approach that recommends a minimum of eight ANC contacts during pregnancy [13]. The 2016 WHO guidelines also emphasise the importance of community-based strategies to promote ANC attendance and improve communication and support. Recommended approaches include encouraging partner and community involvement, improving health literacy, and addressing barriers to ANC access [13].

### 1.3 Barriers to Antenatal Care in Malawi

Malawi’s persistently high maternal mortality rate and low uptake of antenatal care highlight the urgent need to understand women’s perceptions of pregnancy and the factors that influence their access to ANC. ANC often represents a woman’s first point of contact with the health system and provides opportunities to detect complications, promote health behaviours, and link to other services [30].

Multiple, interrelated barriers limit ANC uptake in Malawi and other low- and middle-income countries. Although similar challenges exist across sub-Saharan Africa, the nature and extent of these barriers vary by context [31]. In Malawi, specific factors include distance to health facilities, poverty, gender inequalities, lack of education, and harmful cultural or religious norms [32]. Poor awareness of ANC’s benefits, along with these structural and social constraints, significantly influence whether and when women seek care.

Despite an increase in the number of women giving birth in health facilities and growing global attention to respectful maternity care, mistreatment and abuse during childbirth remain widespread—especially in sub-Saharan Africa [33]. Forms of mistreatment include verbal abuse, non-consented care, breaches of privacy, and physical abuse, all of which violate women’s rights and discourage future engagement with health services. Evidence suggests that women from rural areas and those experiencing labour complications are more likely to face abusive or neglectful treatment from health professionals [34,35].

Contributing factors include poor working conditions, inadequate training, and limited supervision of health personnel, compounded by deeply rooted gender norms and women’s limited decision-making power in their own healthcare [36].

In Malawi, the Ministry of Health has acknowledged these issues and through its Safe Motherhood Initiative has committed to promoting respectful, high-quality care. This includes improving counselling and communication skills among providers and addressing systemic challenges to reduce inequities in service delivery [37,38].

Crucially, women’s previous experiences of care—both positive and negative—strongly influence not only their own willingness to return to facilities but also the broader community’s trust in the health system [39]. If the quality of ANC is perceived as poor, women may delay or avoid care altogether. Conversely, respectful treatment strengthens confidence in facility-based services and promotes greater uptake.

### 1.5 Socio-economic Factors

Socio-economic inequalities remain one of the most significant barriers to accessing antenatal care (ANC) services in Malawi. Distance to health facilities, poverty, and competing responsibilities such as childcare and household duties often prevent women from seeking timely ANC, particularly in rural areas Many women in remote communities walk more than 7 kilometres to reach a health centre, often without food or adequate resources for the journey [40].

Studies have shown that women from wealthier households and urban areas are more likely to initiate ANC earlier and attend more frequently, whereas women in rural settings—despite having comparable needs—face greater obstacles [41]. Additionally, although Malawi has invested in training health professionals, many trained personnel are reluctant to work in rural communities due to limited professional and economic opportunities, resulting in persistent staff shortages in these settings [42,43].

Education also plays a critical role. Women whose partners have higher levels of education are more likely to access skilled ANC services, and household decision-making power further influences maternal health behaviours [44]. Importantly, maternal mortality could be reduced by as much as 70% if all women in sub-Saharan Africa completed at least primary education [45].

Raising women’s socio-economic status is essential for addressing these disparities. Greater access to income, education, and information increases autonomy and enables women to make informed decisions about their health [46]. It also reduces vulnerability to harmful sociocultural practices—such as hiding pregnancies due to fear of witchcraft or delaying care due to stigma associated with being unmarried or young [47–49]. However, Malawi’s reliance on rain-fed agriculture and exposure to climatic shocks has severely constrained economic growth and contributed to household poverty, compounding these challenges [50].

### 1.6 Gender

Gender norms in Malawi are deeply entrenched in patriarchal structures, where men typically act as household decision-makers and gatekeepers of financial and health-related decisions [51]. Although men are often responsible for planning and providing healthcare in the household, their actual participation in antenatal care (ANC) remains low, influenced by social norms, cultural expectations, and fear of HIV testing during ANC visits [52].

The dominance of men in reproductive decision-making limits women’s autonomy, especially when they lack knowledge about maternal health services or need their partner’s approval before attending ANC [53]. Despite growing evidence showing that male involvement improves ANC attendance, birth preparedness, skilled birth and postpartum care, many men continue to perceive maternal and child health as solely the domain of women [54,55].

Studies in Malawi have also highlighted sociocultural practices that reinforce this divide. For example, in some communities, women require an “authorisation letter” from a traditional leader to attend ANC if they become pregnant outside marriage—a practice that further marginalises young or unmarried women [56]. In addition, some districts impose by-laws that penalise women who give birth at home or attend ANC without their husbands, including fines or public shaming [57].

Although these by-laws are often framed as protective and meant to encourage skilled facilitybased care, they can be coercive and may deter already vulnerable women from seeking care [58]. To address this, policies must be sensitive to gendered power dynamics, promote inclusive participation of both partners in reproductive health, and eliminate structural barriers that prevent equitable access to ANC [59,60].

#### Aims and Objectives

This study aimed to explore women’s perceptions of pregnancy and antenatal care (ANC) in Chizenga village, a rural setting in Chikwawa District, Malawi. The objective was to gain an in-depth understanding of how pregnant women living in a resource-limited setting perceive maternal health and ANC, and what barriers influence their utilisation of ANC services.

The study sought to:

1. Understand women’s knowledge of health during pregnancy and perceptions of ANC.
2. Explore socio-cultural and structural barriers that influence the timing and frequency of ANC attendance.
3. Identify opportunities for improving access to and uptake of ANC through community-based or policy-level interventions.

## Materials and Methods

### Study Design

This qualitative interpretive study, conducted in August 2023, was used to explore women’s perceptions of pregnancy and antenatal care in Chizenga village, Chikwawa, Malawi.

Fourteen semi-structured interviews were carried out over a two-week period in the participants’ homes and explored issues around women’s understanding of maintaining health during pregnancy and their perceptions of pregnancy and antenatal care.

### Setting

Chizenga village is situated in Chikwawa, a district in the Southern region of Malawi with a population of 564,684. The population of Chizenga village is approximately 1,000 (61).

Chikwawa is a malaria-endemic area which is prone to climatic shocks and persistent floods and droughts. It is a hotspot for cholera due to poor water and sanitation systems and practices. The district also has a high incidence of challenges affecting children, including child marriages and neglect, with the prevalence of adolescent pregnancies partly influenced by complex legal and cultural norms (62). Chikwawa also has one of the lowest attendance rates for antenatal care services in the first trimester of pregnancy (11%) (63). Barriers to utilising maternal health services in Chikwawa include walking long distances to access health facilities, lack of healthcare personnel, lack of or insufficient items to be used during delivery, long stays, and disrespectful health personnel (64).

Although most women in Chizenga village give birth to their babies at Chizenga Rural Hospital, some still prefer to give birth at home due to the long distance to the hospital. When available, communities use bicycle ambulances to transfer pregnant women quickly from villages in Chikwawa to rural health facilities and district hospitals (65). However, for women who remain at home, births are often assisted by traditional birth attendants (TBAs), most of whom have not received formal training. These attendants are usually older women in the community with experiential knowledge of childbirth but lack the skills or equipment to manage complications. In some cases, women may also give birth entirely on their own, especially if labour begins at night or transport is unavailable.

### Sampling

Purposive sampling was used to select women in their first, second, or third trimester of pregnancy. Only those who were willing to participate and able to give informed consent were included. Women who were not pregnant, who did not reside in Chizenga village, who were not willing to participate, or who were unable to give informed consent were excluded.

Women were recruited between Thursday 1st June 2023 and Saturday 29th July 2023. All participants provided informed consent prior to enrolment in the study. In accordance with ethical requirements, informed consent was obtained verbally and subsequently documented through the signing of a consent form. Each participant either signed her name or, where literacy was limited, provided a thumbprint as confirmation of consent. This process was conducted after the Research Assistant (RA) had explained the purpose, procedures, risks, and benefits of the study in detail, ensuring that participants fully understood their involvement. The RA also served as a witness to the consent process, thereby safeguarding the validity and ethical integrity of participant enrolment.

The women self-selected whether they wished to participate in the study. To reduce bias, the study was geographically restricted to women living close to the centre of Chizenga village. We also screened participants to ensure a range of views and experiences were captured. This included women across different stages of pregnancy, women who were pregnant for the first time (primigravida) as well as those who had one or more children (multigravida), and women from varied marital statuses and educational backgrounds. This approach helped to ensure that the data reflected the diverse experiences of pregnant women living in Chizenga village.

Table 1 outlines the key characteristics of the participants, including marital status, education level, and whether they were primigravida or multigravida.

**Table 1:**
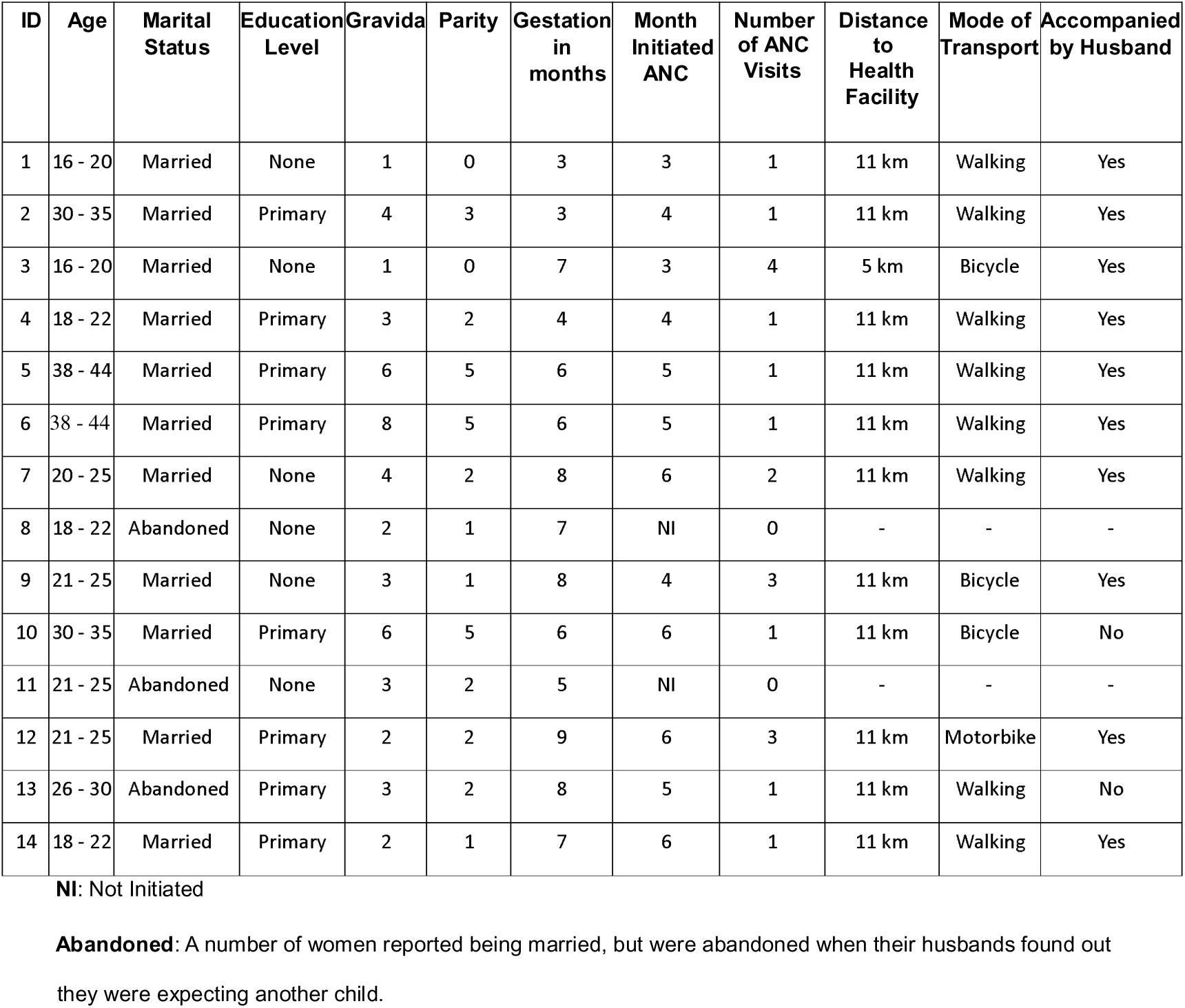
Socio-demographic characteristics of the study sample.

| ID | Age | Marital Status | Education Level | Gravida | Parity | Gestation in months | Month Initiated ANC | Number of ANC Visits | Distance to Health Facility | Mode of Transport | Accompanied by Husband |
| --- | --- | --- | --- | --- | --- | --- | --- | --- | --- | --- | --- |
| 1 | 16 - 20 | Married | None | 1 | 0 | 3 | 3 | 1 | 11 km | Walking | Yes |
| 2 | 30 - 35 | Married | Primary | 4 | 3 | 3 | 4 | 1 | 11 km | Walking | Yes |
| 3 | 16 - 20 | Married | None | 1 | 0 | 7 | 3 | 4 | 5 km | Bicycle | Yes |
| 4 | 18 - 22 | Married | Primary | 3 | 2 | 4 | 4 | 1 | 11 km | Walking | Yes |
| 5 | 38 - 44 | Married | Primary | 6 | 5 | 6 | 5 | 1 | 11 km | Walking | Yes |
| 6 | 38 - 44 | Married | Primary | 8 | 5 | 6 | 5 | 1 | 11 km | Walking | Yes |
| 7 | 20 - 25 | Married | None | 4 | 2 | 8 | 6 | 2 | 11 km | Walking | Yes |
| 8 | 18 - 22 | Abandoned | None | 2 | 1 | 7 | NI | 0 | - | - | - |
| 9 | 21 - 25 | Married | None | 3 | 1 | 8 | 4 | 3 | 11 km | Bicycle | Yes |
| 10 | 30 - 35 | Married | Primary | 6 | 5 | 6 | 6 | 1 | 11 km | Bicycle | No |
| 11 | 21 - 25 | Abandoned | None | 3 | 2 | 5 | NI | 0 | - | - | - |
| 12 | 21 - 25 | Married | Primary | 2 | 2 | 9 | 6 | 3 | 11 km | Motorbike | Yes |
| 13 | 26 - 30 | Abandoned | Primary | 3 | 2 | 8 | 5 | 1 | 11 km | Walking | No |
| 14 | 18 - 22 | Married | Primary | 2 | 1 | 7 | 6 | 1 | 11 km | Walking | Yes |

**NI:** Not Initiated
**Abandoned:** A number of women reported being married, but were abandoned when their husbands found out they were expecting another child.

### Data Collection

Data was collected over a two-week period using semi-structured interviews after obtaining informed, formal written consent. All of the interviews were conducted in Chichewa and took place in the participant’s home. A topic guide was used to keep the interview focused, using open-ended questions and giving the participants greater freedom to describe and discuss their knowledge and perceptions of pregnancy and antenatal care. The interviews were audiorecorded using a Dictaphone and detailed notes were taken during or immediately after the interviews. In addition to in-depth interviews, the principal investigator made observational notes during and after the interviews, which captured non-verbal cues, context, and environmental factors. Field notes were recorded to supplement the interview data and provide further insight into participant behaviour, setting, and researcher reflections. These data contributed to triangulation and supported a richer interpretation of findings.

### Data Analysis

At the end of each day, the data from the semi-structured interviews were reviewed to identify and address any gaps, adapt the topic guide and refine the questioning approach based on emerging insights. Refining the research approach involved adjusting how questions were phrased or sequenced to better elicit rich, relevant responses in subsequent interviews. This iterative process also helped improve the overall quality of data collection (66). The findings were discussed through debriefing sessions between the principal investigator and the research assistant, which allowed the team to capture nuances, clarify meanings, document non-verbal cues, and triangulate data from multiple sources (67).

All interviews were translated into English and transcribed verbatim. Data were analysed using thematic analysis, a method for identifying, coding, and reporting patterns or themes within qualitative data (68). The principal investigator conducted line-by-line coding of the transcripts to generate initial codes, which were then grouped into broader themes and subthemes. Once themes had been developed, transcripts were exported to ATLAS.ti, a qualitative analysis software, to support further coding, content analysis, and interpretation. A sample of transcripts was independently reviewed by the research assistant to support consensus-building, minimise bias, and enhance the validity and accuracy of the findings.

### Quality Assurance

Triangulation was used to increase the credibility of the study which involved answering the research question in several ways; interviews, observation, field note analysis, and substantial description of the interpretation process. Quotations from the data were also used to illustrate and support their interpretation.

### Positionality

The principal investigator is a UK-qualified nurse with over twenty years of experience in maternal and child health, including global health roles across sub-Saharan Africa. At the time of the study, the researcher was undertaking postgraduate studies in Global Health and had previously worked in Malawi on maternal health initiatives. While this experience informed the research focus, it also presented a potential for bias.

To minimise bias, reflexivity was employed throughout the research process. A Malawian research assistant conducted all interviews in Chichewa and supported translation and transcription. The principal investigator maintained a reflective journal to document assumptions, reactions, and observations, thereby enabling critical engagement with the data and awareness of how personal background and positionality may have influenced interpretation.

The study was designed to centre women’s voices and perspectives. The interview guide used open-ended questions to explore participants lived experiences.

## Ethical Considerations and Informed Consent

Ethical approval for the study was obtained from the Liverpool School of Tropical Medicine (UK – Protocol Number: GH23(03)), the College of Medicine Research and Ethics Committee (COMREC – Protocol Number: P.04/23/4064), and the Chikwawa Health Research Coordinating Committee (Malawi).

## Findings

This study explored women’s perceptions of pregnancy and antenatal care (ANC) in Chizenga village, Chikwawa, Malawi. Findings are drawn from fourteen in-depth interviews conducted over a two-week period. The results are presented under six key themes that emerged from the data: women’s understanding of health during pregnancy, dietary beliefs and practices, male involvement in ANC, the perceived role of witchcraft, negative attitudes of healthcare workers, and the impact of distance to healthcare facilities. These themes reflect the complex social, cultural, and structural factors that shape women’s experiences of pregnancy and their engagement with ANC services. Where quotes have been used to understand the voice of the participant, a descriptor has also been provided on marital status, level of education and whether she is pregnant for the first time (primigravida) or is pregnant for at least the second time (multigravida).

## Pregnancy Health Begins with ANC: What Women Know and Expect

The study found that all of the women believed that accessing antenatal care (ANC) was essential to ensuring a healthy pregnancy and safe birth. Participants demonstrated this by describing specific services they expected from ANC visits, such as having their blood pressure, weight, and iron levels checked. Many emphasised the importance of health workers monitoring the baby’s growth and checking for regular movements. Several women also showed knowledge of disease screening during ANC, particularly HIV testing, and spoke about the use of antiretroviral medication to prevent mother-to-child transmission. Others discussed receiving iron tablets, anti-malarial medication, mosquito nets, and access to pain relief. They also identified ANC as a place to receive advice on their pregnancy—particularly antenatal and birth preparedness—and the health of their baby.

‘When we go there (health facility), we will get our weight measured, we stand on the scale. After that we go inside the room and the doctor measures if the baby is kicking or not, after that we get medication that prevents us from malaria. We are also given a mosquito net. We are given HIV screening and vaccinations if we need them.’ (ID 3, Married, no formal education, primigravida)Overall, the participant’s perception of ANC was very positive. All of the women expressed the need to increase education on ANC in Chizenga village to ensure that more women experience a healthy pregnancy and safe birth with skilled health workers at a health facility. However, despite all of the participants understanding the importance of initiating ANC in the first trimester, most women started ANC at around 4 months (16 weeks). All the participants said that they started ANC late due to the long distance they needed to travel to the healthcare facility. Lack of money for transport and food for the journey and husbands not being able to accompany them to their appointment were also reasons the women gave for not attending earlier on in their pregnancy:

‘I have not been to go to an antenatal care appointment yet because I have to go to the hospital so it’s too far. To walk from here (home) to there (healthcare facility) it’s difficult, that’s why I haven’t started yet. I am starting next week. I started antenatal care at four months for my other children too.’ (ID 4, married, primary education, multigravida)All of the women were interested in exploring ways to encourage and increase understanding and awareness of the benefits of ANC in Chizenga village. Each of them believed that better education, better access to health facilities and transport, and increased resources would mean more women would have access to ANC.

### Food Knowledge Without Food Security: Diet

This theme highlights the gap between women’s nutritional knowledge and the harsh economic realities that prevent them from applying it. When the participants were asked what they thought made a pregnancy healthy for both mother and baby, most women talked about the importance of a healthy diet. Several participants also spoke about the food groups healthcare workers had recommended during their ANC appointment.

Several women said they were aware that they should eat more fish, fruit and vegetables; one mother said she had been advised to take vitamin supplements during her previous antenatal appointments. However, most of the women’s responses focused on the fact that they could not vary their diet due to a lack of money and resources. Most of the women interviewed said that they lived on Nsima (a thick porridge made out of maize flour and water), beans and cabbage.

Approximately half of the respondents stated that despite having land, they could not grow enough crops to eat regularly and even when it was possible to sell their grain, there was only enough money to eat twice a day. Other participants said that they could only manage one meal and, at times, would go without food for the whole day:

‘Since my husband went to Mozambique, I don’t get any help, so I don’t have any food. Most of the time I don’t eat. You can see (points to cooking pots with no evidence of fire from the morning) since this morning, I didn’t cook anything, so most of the time I feel dizzy because I am hungry and sometimes, I collapse. So, the real problem is how to get food, I don’t have food.’ (ID 11, abandoned, no formal education, multigravida)

### Alone but Married: Gendered Expectations in ANC Attendance

The participants indicated that the village chiefs actively encouraged men’s involvement in caring for their wives during pregnancy. Men were expected to attend ANC appointments with their wives, which made the women feel that their husbands were also being educated and were able to better understand their wives’ needs than if they had not attended ANC.

Most participants reported that their husbands attended antenatal clinics with them, often describing it as a very positive experience. The women explained that healthcare staff encouraged the involvement of husbands in ANC by giving them preferential treatment:

‘If you are with your husband, you receive better treatment and you are seen first. You are brought to the front of the line ahead of the other women who are attending by themselves. Those without husbands are pushed back and will be the last one to be attended.’ (ID 2, married, primary education, multigravida)

‘We are treated well. When husbands go with their wives they are praised and encouraged to attend each time with their wives so it is a positive experience.’ (ID 4, married, primary education, multigravida)Several participants also disclosed that they felt significantly more supported when their husbands attended ANC with them. They explained that they felt their husbands better understood their needs, the baby’s development, and the importance of birth preparedness following the appointment. The women also reported that, in previous pregnancies, their husbands had attended the hospital when they went into labour. Although they had not been allowed into the delivery room when their wives gave birth, they were permitted to wait outside and were able to support their wives on the journey home.

Two of the participants said that although their husbands are supportive and will accompany them to ANC, they do not feel comfortable as a man going inside to the appointment. Other women with more than one child said that their husbands were needed at home to care for their other children and could not escort them to the hospital. Despite being married, as these women arrived at the hospital alone, they were treated differently from women attending with their husbandsWhen asked what happens at the hospital when the women go alone, several women explained: ‘You get the lowest priority.’

(ID 7, married, no formal education, multigravida)Regardless of marital status, unaccompanied women are kept waiting or ignored ‘My husband is in Mozambique so I walk alone to the hospital. Those who attend with their husbands receive better treatment. I get treated very, very late because the doctors think I’m not married and that maybe the pregnancy is out of wedlock so I am not treated like other married women.’ (ID 8, abandoned, no formal education, multigravida)

The treatment pregnant women received from health workers at the hospital significantly influenced how women felt about attending ANC. Therefore, it poses a potential barrier to ANC as other women may choose not to attend to avoid experiencing negative treatment [44,45].

### Seen and Unseen Threats: Navigating Pregnancy in a Context of Witchcraft Beliefs

Only one of the women spoke about beliefs around witchcraft during the in-depth interviews. The participant explained that several years ago, most women in the village did not attend ANC. She said that because women had not gone to ANC and had chosen to give birth at home, there had been considerable maternal and neonatal deaths in the village. However, at the time, the woman explained that everyone believed that the deaths were not due to poor ANC attendance or lack of skilled health personnelduring childbirth but that it was actually due to witchcraft.

The participant said that in response to the high numbers of maternal and neonatal deaths in the village, the village chiefs had called a meeting to encourage all women to attend ANC to prevent further deaths in the community. The woman explained that the chiefs had made it obligatory for all women in Chizenga village to attend antenatal appointments with their husbands and to give birth at a healthcare facility.

One participant believed that since changes had been implemented, there had been no further maternal deaths in the village, which she attributed to more women attending antenatal care.

While this reflects her personal perception and optimism about the impact of ANC, maternal mortality data specific to the village were not available to confirm this. However, the same participant acknowledged that, despite increasing awareness of the benefits of ANC and facility-based births, some women still feared that witchcraft could be used against them during pregnancy. As a result, they concealed their pregnancies until the second or third trimester and sometimes travelled to the hospital alone at night to avoid being bewitched.

‘If fewer people know you are going to the hospital, there is less chance of the mother and baby dying due to witchcraft. Once you are at the hospital you are safe because witchcraft doesn’t work at the hospital.’ (ID 5, married, primary education, multigravida)

### Respect Shapes Access: How Attitudes Influence ANC Uptake – Negative Attitude of Healthcare Workers

Most of the participants in this study described having a positive experience of healthcare workers during ANC appointments at the hospital. The women reported that they felt welcomed and explained that they were treated respectfully during their ANC visits at the healthcare facility. However, there were exceptions, and not all of the participants felt that they had been treated respectfully during their ANC appointment as shown below:

‘If women have forgotten the date of their appointment or you attend on a different date, you get shouted at.’ (ID 3, married, no formal education, primigravida)

‘If I have no money for food and no husband, I receive different treatment from the healthcare staff, so it’s better to stay here (at home) as I don’t want to be treated badly. Being treated differently feels painful.’ (ID 11, abandoned, no formal education, multigravida)Although most women had either experienced or were aware of others who had experienced negative attitudes from healthcare workers during their ANC appointment at the hospital, they all believed attending ANC during pregnancy was important and wanted to continue despite the disrespectful treatment at times.

### Beyond Distance: Women’s Accounts of Accessing Antenatal Services

Distance significantly impacted when women accessed ANC as their nearest health facility was 11 km from Chizenga village. Most women reported starting antenatal care late due to the distance they would need to travel to the hospital and lack of transportation. All of the women said that they walked to the hospital and back unless they could borrow a bicycle from someone in the village. Some of the women chose to hire a bike and were taken to their ANC appointment by their husbands. They did this so they did not have to be away from their older children for too long.

Women who could borrow or rent a bicycle and attend their ANC visits with their husbands were able to complete the journey to the health facility and back within five hours. Some participants also mentioned the availability of bicycle taxis, but explained that these were not always reliable or available when needed. Renting a bicycle or hiring a bicycle taxi from Chizenga to the nearest healthcare facility and back cost around 4000 MWK ($3.70), a costly option given that 70% of the population in Malawi lives on less than 2300 MWK ($1.75) a day [61].

‘Borrowing a bike can be expensive for us because when we have a bicycle and it breaks down, we have to pay. The bicycle we used last time broke down three times and we had to repair it. We also have to buy food if we get hungry.’ (ID 3, married, no formal education, primigravida)

Women who were unable to borrow or rent a bicycle reported that the journey to and from the hospital took the entire day, as they had to walk long distances and then wait for hours to be seen—often despite arriving early:

‘I need to start walking at 7 in the morning so that I can come back from the hospital to look after my other children in the afternoon.’ (ID 5, married, primary education, multigravida)

Despite the women’s challenges, all of them were keen to access ANC healthcare despite the long distance to the health centre and hospital. Most of the women felt that the hospital was the only place they could access good advice on their health and the baby’s health. All participants indicated that there was a lack of support for pregnant women in the village, with only two or three saying they would seek advice from their mothers if needed. When asked if all of the women in the village access ANC during pregnancy, most of the women explained that while everyone should attend, there are women who:

‘Don’t want to walk long distances or don’t have money to buy food for the journey, so they stay home.’ (ID 6, married, primary education, multigravida)

The participants believed that some women in the community did not attend ANC due to “laziness”) or “ignorance” (ID 12, married, primary education, multigravida). They explained that these women either lacked awareness of the benefits of ANC or failed to prioritise their health and that of the baby. Some participants expressed frustration that, despite the availability of services, there were still women who chose not to attend unless they were already unwell or in labour. Others felt that this behaviour was rooted in limited understanding and called for more education within the community to raise awareness about the importance of ANC:

We need community health workers to educate women to tell them how important it is and what services are offered.’ (ID 2, married, primary education, multigravida)

All of the women spoke openly about the need for community health workers to increase education on the importance of antenatal care (ANC) during pregnancy, and to raise awareness about the services available. They also emphasised the need for improved transport to enable more women to deliver at a health facility. The participants believed that ANC attendance and health-seeking behaviours would improve further if a health facility were established within Chizenga village.

## Discussion

This study explored women’s perceptions of pregnancy and antenatal care in Chizenga village, Chikwawa, Malawi. The study found that despite many challenges, all of the women in this study valued ANC and demonstrated a determination to do their best and overcome the challenges of accessing ANC to improve health outcomes for themselves and their babies during pregnancy.

This study found that all participants believed it was essential to access ANC during pregnancy and showed an awareness of the importance of diet, monitoring the baby’s growth, monitoring blood pressure and haemoglobin levels, accessing appropriate screening such as HIV and giving birth at a healthcare facility. However, despite the participants’ understanding of the benefits of antenatal care during pregnancy, they lacked knowledge of the importance of early ANC booking.

Consistent with previous studies, the participants were also unaware of the Focused Antenatal Care (FANC) program that recommends at least four ANC appointments for women with uncomplicated pregnancies, with most of the participants initiating ANC at around 16 weeks [62–65].

Multiparity did not influence the early initiation of ANC. Similarly, the findings in this study are similar to previous studies where higher parity is associated with late initiation of ANC. In this study, women with four or more previous births typically initiated antenatal care between five and six months of pregnancy. This delayed uptake is concerning, particularly given the long distance to the healthcare facility, as higher parity is associated with increased risk of complications such as postpartum haemorrhage and obstructed labour [66–68].

These findings suggest that clearer guidance is needed to support women—particularly those with higher parity—in understanding the importance of initiating antenatal care during the first trimester. Improved counselling during community outreach or early contact with health workers may help address misconceptions and encourage timely attendance. Evidence from a study in South Ethiopia supports this, showing that women who did not receive advice on when to begin ANC were three times more likely to initiate care late compared to those who received such guidance [69–71].

However, this study found that ANC initiation was not determined solely by parity but by other factors such as the long distance to the healthcare facility, lack of money and food for the journey and husbands not being able to accompany women to their appointments. These findings are also consistent with findings from previous studies [72–76]. All participants recognised the challenges in accessing ANC and identified how they thought they could overcome some of the challenges. The women expressed a need for community health workers’ presence in Chizenga village. They believed that by improving access to education and explaining the benefits of ANC to women in Chizenga village, more women would make better decisions regarding their health during pregnancy, attend ANC and choose facilitybased care for childbirth.

Evidence shows that interventions involving community health workers can increase knowledge and awareness in rural and under-served communities through community discussions. These approaches also help strengthen links between communities and health facilities, leading to greater utilisation of facility-based maternal care [77]. Community health workers live within the community, so they better understand local beliefs and practices that may influence a woman’s knowledge and attitudes toward antenatal care. This insight can help shape strategies to promote early ANC attendance, which is vital for improving maternal and newborn health outcomes.[78–80]. One study in rural Malawi found that community health worker interventions can significantly increase the utilisation of ANC. The intervention used community health workers to identify pregnant women and link them to ANC at healthcare facilities. This intervention tripled the proportion of women starting ANC in the first trimester [81].

Overall, this study found that participants spoke passionately about the importance of attending ANC and giving birth at a healthcare facility. However, several women explained that women also go because if they do not, they will be punished by the village chiefs.

Village chiefs play an important role in encouraging antenatal care (ANC) and facility-based childbirth. However, while their support has helped increase the number of women attending health facilities with skilled birth attendants, some bylaws are enforced in a coercive way, which may negatively affect how women view ANC and giving birth at a facility [82–84].Several of the participants used the words “encourage” and “advise” when suggesting how to reach out to other women in the village. Evidence suggests that local community members—including family members, neighbours, and women’s groups—play a crucial role in supporting pregnant women. In particular, strengthening women’s groups has been shown to improve ANC uptake and reduce maternal and neonatal mortality in rural settings [77].

However, although Aboagye et al. (2023) argue that promoting women’s empowerment programs without intensive improvements in women’s socio-economic status would be ineffective, they contend that combining empowerment with active improvements in socioeconomic status can encourage women to improve health-seeking behaviours and ANC uptake in rural communities [85]. Unfortunately, despite how determined the participants were to increase knowledge and awareness of ANC in their community, the women felt they needed support.

In rural Malawian communities, where decision-making often follows hierarchical structures led by chiefs and elders, top-down policy implementation may be more effective than bottomup empowerment approaches. Walsh et al. (2018) argue that utilitarian, authority-driven models are more aligned with local expectations and social norms, and may therefore be more successful in influencing health behaviours [84]. However, the use of coercive measures— such as imposing penalties on vulnerable women in resource-poor settings—risks reinforcing health, economic, and gender inequities within these rural communities..

## Strengths and Limitations

This study adds value to the existing literature by showing that positive perceptions of pregnancy and antenatal care from women living in a resource-poor setting exist.

The study shows that women value ANC and want to increase knowledge and awareness of its benefits, not only for themselves but also for other women in their community. A key strength of this study is that data were collected from pregnant women at varying stages of pregnancy, including some who had experienced miscarriage or neonatal loss. These experiences added depth to the findings and contributed to the richness and quality of the data. Using a qualitative approach also allowed the researchers to capture participants’ perceptions and experiences in their natural setting.

Nonetheless, the study had limitations. Firstly, it focused on women over the age of eighteen years. Given the number of adolescent pregnancies reported in Malawi, future studies could also include adolescent women’s perceptions of pregnancy and ANC. Furthermore, the study was conducted in one village and may not reflect the understanding and perceptions of women from other villages in Chikwawa. Finally, only women were included in the study. Given men’s increasing involvement in ANC, future research could explore men’s understanding and perspectives of pregnancy and antenatal care, including the role of village chiefs in influencing care-seeking behaviours. It would also be valuable to include the perspectives of healthcare workers, whose insights could help triangulate the findings and provide a more comprehensive understanding of the barriers and facilitators to ANC uptake in this setting [86].

## Conclusion and Recommendations

This study demonstrates that, despite considerable barriers, women in Chizenga were determined to access facility-based ANC, recognising its importance in ensuring healthy pregnancy outcomes. However, their ability to do so was often constrained by multiple challenges. These included long distances to health facilities, limited financial resources, and competing domestic responsibilities, all shaped by the wider socio-economic context in which the women lived.

The findings show that the participants value ANC at a healthcare facility. However, consistent with the literature, their socio-economic context makes access to ANC challenging. Despite the many challenges, the participants demonstrated remarkable resilience. They expressed a strong desire to support other women in the community and highlighted the importance of improving access to antenatal care through practical, context-specific strategies. These included conducting outreach and sensitisation campaigns, introducing community health workers to strengthen antenatal health education, and increasing social and logistical support for pregnant women at the community level.

The 2016 WHO recommendations on antenatal care for a positive pregnancy experience highlight the importance of community-based interventions to improve communication and support. Furthermore, the recommendations suggest that interventions that improve the dialogue around barriers and enablers to utilising ANC services, maintaining health during pregnancy, and increasing support for women and their partners during pregnancy could significantly improve ANC uptake and quality of care [87].Furthermore, research suggests that eight or more contacts for antenatal care can reduce perinatal deaths by up to eight per thousand births compared to four visits [87]. However, given the number of women who fail to complete four ANC visits in rural communities in Malawi, we need to consider how health systems can be strengthened to meet the WHO guidance and support more women in accessing ANC.

Until now, women living in rural communities have been expected to walk long distances when they are heavily pregnant, in labour, or after giving birth. The participants in Chizenga village were prepared to walk over twenty-two kilometres to attend ANC, with very little to eat or drink and to a place where they may be treated disrespectfully. This shows how valuable the service is to them and demonstrates their commitment to improving health outcomes for themselves and their children. Therefore, governments must recognise this commitment and respond by ensuring that all women, regardless of socio-economic, educational or marital status, have equal access to respectful ANC and the highest attainable health outcomes in pregnancy for the mother and child.

Future strategies to increase the uptake of ANC in rural communities should be designed creatively and context-specific to reflect the challenges women in resource-poor settings face. Strategies should aim to improve women’s educational and socio-economic status to reduce inequalities in access to ANC and other maternal healthcare services. Evidence shows that maternal mortality could decrease by up to 70% if all women in sub-Saharan Africa completed primary education [88]. Moreover, women from wealthier households and those with higher educational attainment are more likely to initiate ANC earlier and attend more frequently, highlighting the urgent need to address structural inequalities in maternal healthcare access [23,88,28]. Recent studies further corroborate the critical role of women’s education and economic status in ANC utilisation. Adedokun et al. (2022) found that higher educational attainment and improved economic conditions significantly increase the likelihood of adequate ANC service use [26]. Similarly, Aboagye et al. and Tessema et al. identified wealth and education as key determinants influencing early initiation and frequency of ANC visits [44,89]. These findings underscore the need to implement strategies that enhance women’s educational and economic opportunities to reduce disparities in maternal health outcomes.

The provision of ANC services could also be expanded to reduce the distance and costs associated with ANC attendance. This could be accomplished by deploying community health workers and midwives to deliver effective and appropriate sensitisation, maternal health campaigns, and mobile clinics in remote communities. Evidence shows that task shifting—delegating elements of maternal healthcare to trained non-physician health workers such as community health workers—can significantly improve access to ANC and continuity of care in low-resource settings [89,87]. Community health workers could also help to support and maintain connections between vulnerable populations and healthcare providers.

Healthcare professionals could consider redefining ANC spaces so that men feel more included, and the idea that ANC is a woman’s domain begins to fade. This study further recommends that health managers ensure staff training and supervision supports sustained care and respect for all women attending ANC, regardless of socio-economic, educational, and marital status.

## Data Availability

Due to the sensitive nature of the data and to protect participant confidentiality, the full dataset cannot be made publicly available. However, anonymised data may be shared upon reasonable request and with appropriate institutional ethical approvals.

## Acknowledgements

The author gratefully acknowledges **Dr Terry Kana** of the Liverpool School of Tropical Medicine for her academic guidance, invaluable encouragement, and steadfast mentorship throughout the study. Thanks, are also extended to **Deborah Nyirenda**, in-country supervisor in Malawi, for her thoughtful direction and meaningful contributions during fieldwork.

Special thanks to **Felia Malola**, research assistant, whose dedication, vision, and tireless commitment were integral to this study. Felia’s unwavering efforts to improve health for women, men, and children in Malawi were deeply respected. Her passing shortly after the study’s completion is a profound loss; her compassion and devotion will be remembered always.

The author also wishes to thank **Lloyd Malola** for his generous hospitality, cultural insight, and enduring support, as well as the participants and village leaders of **Chizenga** for their trust, time, and openness in sharing their experiences.

Finally, heartfelt appreciation is extended to the author’s **husband, children, and parents**, whose love, encouragement, and unfailing belief were a constant source of strength and purpose throughout this journey.

## References

1. National Statistical Office (NSO) [Malawi] and ICF. Malawi Demographic and Health Survey 2015-16. Zomba, Malawi, and Rockville, Maryland, USA: NSO and ICF; 2017.

2. Kuuire VZ, Kangmennaang J, Atuoye KN, Antabe R, Boamah SA, Vercillo S, et al. Timing and utilisation of antenatal care service in Nigeria and Malawi. Glob Public Health. 2017;12(6):711–27.

3. Kifle D, Azale T, Gelaw YA, Yayehirad AM. Maternal health care service seeking behaviours and associated factors among women in rural Haramaya District, Eastern Ethiopia: a triangulated community-based cross-sectional study. Reprod Health. 2017;14(1):6.

4. Mathias CT, Mianda S, Ginindza TG. Facilitating factors and barriers to accessibility and utilization of kangaroo mother care service among parents of low-birth-weight infants in Mangochi District, Malawi: a qualitative study. BMC Pediatr. 2020;20(1):355.

5. Castleberry A, Nolen A. Thematic analysis of qualitative research data: Is it as easy as it sounds? Curr Pharm Teach Learn. 2018;10(6):807–15.

6. World Health Organization. Definition of Skilled Health Personnel Providing Care during Childbirth: the 2018 Joint Statement by WHO, UNFPA, UNICEF, ICM, ICN, FIGO and IPA. Geneva: WHO; 2018.

7. World Health Organization. Maternal Mortality: Key Facts [Internet]. Geneva: WHO; 2025. Available from: https://www.who.int/news-room/fact-sheets/detail/maternalmortality

8. Geller SE, Koch AR, Garland CE, MacDonald JE, Storey F, Lawton B. A global view of severe maternal morbidity: moving beyond maternal mortality. Reprod Health. 2018;15(Suppl 1):98.

9. Moyer CA, Lawrence ER, Beyuo TK, Tuuli MG, Oppong SA. Stalled progress in reducing maternal mortality globally: what next? Lancet. 2023;401(10382):1060–2.

10. World Health Organization. Trends in maternal mortality 2000 to 2020: estimates by WHO, UNICEF, UNFPA, World Bank Group and UNDESA/Population Division. Geneva: WHO; 2023.

11. United Nations Children’s Fund (UNICEF). Trends in maternal mortality 2000 to 2020 [Internet]. 2023. Available from: https://data.unicef.org/resources/trends-inmaternal-mortality-2000-to-2020/

12. World Health Organization. Maternal Mortality: Key Facts [Internet]. Geneva: WHO; 2023 [cited 2023 Feb 22].

13. Dickson KS, Okyere J, Ahinkorah BO, Seidu AA, Salihu T, Bediako V, et al. Skilled antenatal care services utilisation in sub-Saharan Africa: a pooled analysis of demographic and health surveys from 32 countries. BMC Pregnancy Childbirth. 2022;22(1):831.

14. Duodu PA, Bayuo J, Mensah JA, Livingstone AP, Holmes FA, Dzomeku VM, et al. Trends in antenatal care visits and associated factors in Ghana from 2006 to 2018. BMC Pregnancy Childbirth. 2022;22(1):59.

15. United Nations Children’s Fund (UNICEF). Chikwawa district pilots child-centred development planning [Internet]. 2021. Available from: https://www.unicef.org/malawi/stories/chikwawa-district-pilots-child-centereddevelopment-planning

16. Baluwa PC. Utilization of antenatal care services in the first trimester of pregnancy: Analysis of facility-based barriers and potential interventions in Chikwawa district [Master’s thesis]. Kamuzu University of Health Sciences; 2022.

17. Kambala C, Morse T, Masangwi S, Mitunda P. Barriers to maternal health service use in Chikhwawa, Southern Malawi. Malawi Med J. 2011;23(1):1–5.

18. Ministry of Health (Malawi). Presidential Initiative on Maternal Health & Safe Motherhood [Internet]. 2022. Available from: https://www.health.gov.mw/presidential-initiative-on-martenal-health-safemotherhood/

19. Busetto L, Wick W, Gumbinger C. How to use and assess qualitative research methods. Neurol Res Pract. 2020;2:14.

20. McMahon SA, Winch PJ. Systematic debriefing after qualitative encounters: an essential analysis step in applied qualitative research. BMJ Glob Health. 2018;3(5):e000837.

21. Mchenga M, Burger R, von Fintel D. Examining the impact of WHO’s Focused Antenatal Care policy on early access, underutilisation and quality of antenatal care services in Malawi: a retrospective study. BMC Health Serv Res. 2019;19(1):295.

22. Okedo-Alex IN, Akamike IC, Ezeanosike OB, Chigozie JU. Determinants of antenatal care utilisation in sub-Saharan Africa: a systematic review. BMJ Open. 2019;9(10):e031890.

23. Tekelab T, Chojenta C, Smith R, Loxton D. Factors affecting utilization of antenatal care in Ethiopia: A systematic review and meta-analysis. PLoS One. 2019;14(4):e0214848.

24. Chimatiro C, Hajison P, Chipeta E, Muula A. Understanding barriers preventing pregnant women from starting antenatal clinic in the first trimester of pregnancy in Ntcheu District-Malawi. Reprod Health. 2018;15(1):1–8.

25. Maharaj R, Mohammadnezhad M. Perception of pregnant women towards early antenatal visit in Fiji: a qualitative study. BMC Pregnancy Childbirth. 2022;22(1):111.

26. Adedokun ST, Yaya S. Correlates of antenatal care utilization among women of reproductive age in sub-Saharan Africa: evidence from multinomial analysis of demographic and health surveys (2010–2018) from 31 countries. Arch Public Health. 2020;78:134.

27. Wolde F, Mulaw Z, Zena T, Biadgo B, Limenih MA. Determinants of late initiation for antenatal care follow up: the case of northern Ethiopian pregnant women. BMC Res Notes. 2018;11(1):837.

28. Seidu AA, Ameyaw EK, Sambah F, Baatiema L, Oduro JK, Budu E, et al. Type of occupation and early antenatal care visit among women in sub-Saharan Africa. BMC Public Health. 2022;22(1):1118.

29. de Masi S, Bucagu M, Tunçalp Ö, Peña-Rosas JP, Lawrie T, Oladapo OT, et al. Integrated person-centered health care for all women during pregnancy: implementing WHO recommendations on antenatal care for a positive pregnancy experience. Glob Health Sci Pract. 2017;5(2):197–201.

30. Gebremeskel F, Dibaba Y, Admassu B. Timing of first antenatal care attendance and associated factors among pregnant women in Arba Minch Town and Arba Minch District, Gamo Gofa Zone, South Ethiopia. J Environ Public Health. 2015;2015:971506

31. Nyando M, Makombe D, Mboma A, Mwakilama E, Nyirenda L. Exploring perceptions of pregnant women on antenatal care visit during their first trimester at Area 25 Health Centre in Lilongwe, Malawi – A qualitative study. Res Square. 2023. 10.21203/rs.3.rs-3140322/v1

32. Mgata S, Maluka SO. Factors for late initiation of antenatal care in Dar es Salaam, Tanzania: a qualitative study. BMC Pregnancy Childbirth. 2019;19(1):1–9.

33. Mamba KC, Muula AS, Stones W. Facility-imposed barriers to early utilization of focused antenatal care services in Mangochi District, Malawi – a mixed methods assessment. BMC Pregnancy Childbirth. 2017;17(1):44.

34. Manda-Taylor L, Sealy DA, Roberts J. Factors associated with delayed antenatal care attendance in Malawi: Results from a qualitative study. Med J Zambia. 2017;44(1):17–25.

35. Moller AB, Petzold M, Chou D, Say L. Early antenatal care visit: a systematic analysis of regional and global levels and trends of coverage from 1990 to 2013. Lancet Glob Health. 2017;5(10):977–83.

36. Pindani M, Chilinda I, Botha J, Chorwe-Sungani G. Exploring community support on safe motherhood: a case of Lilongwe District, Malawi. Afr J Prim Health Care Fam Med. 2021;13(1):a2972.

37. Thapa DK, Acharya K, Karki A, Cleary M. Health facility readiness to provide antenatal care (ANC) and non-communicable disease (NCD) services in Nepal and Bangladesh: analysis of facility-based surveys. PLoS One. 2023;18(3):e0283044.

38. Kok MC, Vallières F, Tulloch O, Kumar MB, Kea AZ, Karuga R, et al. Does supportive supervision enhance community health worker motivation? A mixedmethods study in four African countries. Health Policy Plan. 2018;33(9):988– 98.

39. Djellouli N, Mann S, Nambiar B, Meireles DM, Barros H, Bocoum FY, et al. Improving postpartum care delivery and uptake by implementing context-specific interventions in four countries in Africa: a realist evaluation of the Missed Opportunities in Maternal and Infant Health (MOMI). BMJ Glob Health. 2017;2(4):e000408.

40. Kachimanga C, Dunbar EL, Watson S, Cundale K, Makungwa H, Wroe EB, et al. Increasing utilisation of perinatal services: estimating the impact of a community health worker program in Neno, Malawi. BMC Pregnancy Childbirth. 2020;20(1):22.

41. Lodenstein E, Ingemann C, Molenaar JM, Dieleman M, Broerse JEW. Informal social accountability in maternal health service delivery: A study in Northern Malawi. PLoS One. 2018;13(6):e0195671.

42. Sharma BB, Jones L, Loxton DJ, Booth D, Smith R. Systematic review of community participation interventions to improve maternal health outcomes in rural South Asia. BMC Pregnancy Childbirth. 2018;18(1):327.

43. Walsh A, Matthews A, Manda-Taylor L, Brugha R, Mwale D, Phiri T, et al. The role of the traditional leader in implementing maternal, newborn and child health policy in Malawi. Health Policy Plan. 2018;33(8):879–87.

44. Aboagye RG, Okyere J, Seidu AA, Ahinkorah BO, Budu E, Yaya S. Does women’s empowerment and socio-economic status predict adequacy of antenatal care in sub-Saharan Africa? Int Health. 2023. 10.1093/inthealth/ihad016

45. World Health Organization. WHO recommendations on antenatal care for a positive pregnancy experience. Geneva: WHO; 2016.

46. Ng’ambi W, Mphande M, Banda L, Phiri D. Examining educational attainment and maternal health service utilization in sub-Saharan Africa. Int J MCH AIDS. 2022;11(2):e516.

47. Sserwanja Q, Kawuki J, Kim JH. Socioeconomic factors associated with utilization of maternal health services in sub-Saharan Africa: a multilevel analysis. BMC Public Health. 2022;22(1):1122.

48. Joshi C, Torvaldsen S, Hodgson R, Hayen A. Factors associated with the use and quality of antenatal care in Nepal: a population-based study using the demographic and health survey data. BMC Pregnancy Childbirth. 2014;14:94.

49. Machira K, Palamuleni ME. Women’s knowledge and attitudes towards maternal health care services in Malawi. Afr Health Sci. 2018;18(3):819–27.

50. Kazanga I, Munthali AC, Malata A. Reducing maternal mortality in Malawi: Is education the solution? Malawi Med J. 2019;31(2):131–4.

51. Amponsah MO, Dimbuene ZT, Udofia EA. Do antenatal care providers deliver health education messages? A qualitative study in two Ghanaian hospitals. BMC Health Serv Res. 2021;21(1):1016.

52. Lungu E, Darkwah S, Kafulafula U. Health education during antenatal care: a case study of the provision and effectiveness in Malawi. Int J Afr Nurs Sci. 2023;18:100494.

53. Dahab R, Sakellariou D. Barriers to accessing maternal care in low income countries in Africa: a systematic review. Int J Environ Res Public Health. 2020;17(12):4292.

54. Diamond-Smith N, Sudhinaraset M, Montagu D, Brizuela V, Sharma J. Women’s experiences of disrespect and abuse during childbirth in health facilities in Malawi: a qualitative study. Reprod Health. 2022;19(1):107.

55. Ali NA, Osman MM, Abbaker AO, Adam AM, Adam MS. Prevalence and factors associated with disrespect and abuse during facility-based childbirth in Central Sudan. BMC Pregnancy Childbirth. 2020;20(1):1–7.

56. Kassa ZY, Tsegaye B, Abeje A. Disrespect and abuse of women during childbirth in public health facilities in Addis Ababa, Ethiopia. BMC Pregnancy Childbirth. 2020;20(1):658.

57. Gebeyehu S, Hailu D, Seid SS, Muche A, Melketsedik ZT. Factors associated with disrespect and abuse during childbirth among women delivering at a teaching hospital in Southwest Ethiopia. BMC Pregnancy Childbirth. 2023;23(1):1–10.

58. Ahmed S, Creanga AA, Gillespie DG, Tsui AO. Economic status, education and empowerment: implications for maternal health service utilization in developing countries. PLoS One. 2023;18(3):e0267983.

59. Merriel A, Lisy K, Biswas A, Lawrence K, Vogel JP. Respectful maternity care for women experiencing stillbirth: a systematic review. BJOG. 2021;128(10):1752–63.

60. Mannava P, Durrant K, Fisher J, Chersich M, Luchters S. Attitudes and behaviours of maternal health care providers in interactions with clients: a systematic review. Glob Health. 2015;11:36.

61. Clark C, Ferguson L, Shakya H, Harper M. Influence of gender norms and decisionmaking power on women’s access to health services in Malawi. Reprod Health. 2020;17(1):163.

62. Kohi TW, Mselle LT, Dol J, Kothari A, de Mello MC. Male involvement in reproductive and maternal health care: A qualitative study among community members in rural Tanzania. Reprod Health. 2018;15(1):18.

63. Sakala B, Kazanga I, Tchongwe P. Involving men in antenatal care in Malawi: the perspectives of health providers. J Midwifery Reprod Health. 2021;9(3):2755–63.

64. Tokhi M, Comrie-Thomson L, Davis J, Portela A, Chersich M, Luchters S. Involving men to improve maternal and newborn health: A systematic review of the effectiveness of interventions. PLoS One. 2018;13(1):e0191620.

65. Suandi D, Williams P, Bhattacharya S. Does male involvement improve maternal and newborn health outcomes in developing countries? A systematic review. J Obstet Gynaecol. 2019;39(7):926–32.

66. Smith J, Portela A, Marston C. Improving respectful maternity care in health facilities: a conceptual mapping exercise. Reprod Health. 2022;19(1):11.

67. Le Meur N. Are by-laws a barrier to maternal healthcare in Malawi? A policy brief. Evidence for Action. 2015. https://www.evidence4action.net/resources/policy-briefare-laws-barrier-maternal-healthcare-malawi

68. United Nations Population Fund (UNFPA). National Sexual and Reproductive Health and Rights Policy. Lilongwe: Ministry of Health, Malawi; 2017.

69. Mchenga M, Burger R, Von Fintel D. Examining the impact of WHO’s Focused Antenatal Care policy on early access, underutilisation and quality of antenatal care services in Malawi: a retrospective study. BMC Health Serv Res. 2019;19(1):295.

70. United Nations Children’s Fund (UNICEF). Chikwawa district pilots child-centred development planning. 2021. https://www.unicef.org/malawi/stories/chikwawadistrict-pilots-child-centered-development-planning

71. Baluwa PC. Utilisation of antenatal care services in the first trimester of pregnancy: Analysis of facility-based barriers and potential interventions in Chikwawa district. Kamuzu University of Health Sciences; 2022.

72. Kambala C, Morse T, Masangwi S, Mitunda P. Barriers to maternal health service use in Chikhwawa, Southern Malawi. Malawi Med J. 2011;23(1):1–5.

73. Ministry of Health (Malawi). Presidential Initiative on Maternal Health & Safe Motherhood. 2022. https://www.health.gov.mw/presidential-initiative-on-martenalhealth-safe-motherhood/

74. Busetto L, Wick W, Gumbinger C. How to use and assess qualitative research methods. Neurol Res Pract. 2020;2:14.

75. McMahon SA, Winch PJ. Systematic debriefing after qualitative encounters: an essential analysis step in applied qualitative research. BMJ Glob Health. 2018;3(5):e000837.

76. Maharaj R, Mohammadnezhad M. Perception of pregnant women towards early antenatal visit in Fiji: a qualitative study. BMC Pregnancy Childbirth. 2022;22(1):111.

77. Adedokun ST, Yaya S. Correlates of antenatal care utilization among women of reproductive age in sub-Saharan Africa: evidence from multinomial analysis of demographic and health surveys (2010–2018) from 31 countries. Arch Public Health. 2020;78(1):134.

78. Wolde F, Mulaw Z, Zena T, Biadgo B, Limenih MA. Determinants of late initiation for antenatal care follow up: the case of northern Ethiopian pregnant women. BMC Res Notes. 2018;11(1):837.

79. Seidu AA, Ameyaw EK, Sambah F, Baatiema L, Oduro JK, Budu E, et al. Type of occupation and early antenatal care visit among women in sub-Saharan Africa. BMC Public Health. 2022;22(1):1118.

80. De Masi S, Bucagu M, Tunçalp Ö, Peña-Rosas JP, Lawrie T, Oladapo OT, et al. Integrated person-centered health care for all women during pregnancy: Implementing World Health Organization recommendations on antenatal care for a positive pregnancy experience. Glob Health Sci Pract. 2017;5(2):197–201.

81. Gebremeskel F, Dibaba Y, Admassu B. Timing of first antenatal care attendance and associated factors among pregnant women in Arba Minch Town and Arba Minch District, Gamo Gofa Zone, South Ethiopia. J Environ Public Health. 2015;2015:1–7.

82. Ng’ambi W, Banda L, Mphande M. Women’s education and ANC service utilisation in Malawi: a population-based study. Int J MCH AIDS. 2022;11(2):e516.

83. Adedokun ST, Yaya S. Exploring the link between maternal education and maternal healthcare utilisation in sub-Saharan Africa: a multilevel analysis. BMC Public Health. 2025;25(1):15.

84. Tessema GA, Mahmood MA, Gomersall JS, Assefa Y, Gebrekidan KG, Zemedu TG, et al. Improving maternal health services in sub-Saharan Africa: a review of key predictors and effective interventions. Lancet Glob Health. 2023;11(2):e168–78.

85. World Bank Group. World Development Indicators. 2023. https://databank.worldbank.org/source/world-development-indicators

86. World Health Organization. Definition of skilled health personnel providing care during childbirth: the 2018 joint statement by WHO, UNFPA, UNICEF, ICM, ICN, FIGO and IPA. Geneva: WHO; 2018.

87. World Health Organization. Trends in maternal mortality 2000 to 2020. Geneva: WHO; 2023.

88. World Health Organization. Maternal Mortality Fact Sheet. Geneva: WHO; 2023. https://www.who.int/news-room/fact-sheets/detail/maternal-mortality

89. World Health Organization. WHO recommendations on antenatal care for a positive pregnancy experience. Geneva: WHO; 2016. https://www.who.int/publications/i/item/9789241549912

